# The South-East Asian Transgender Health Cohort: A Protocol for a Prospective Observational Study

**DOI:** 10.1101/2025.10.16.25336946

**Authors:** Rena Janamnuaysook, Tanabodee Payuha, Somporn Saiweaw, Thanh Le, Ronivin Pagtakhan, Pearparn Pumphosuwan, Tidarat Amatsombat, Vitchayada Sripoonbun, Nichaphat Watthanakittikul, Theeranat Sangpasert, Kasiphum Mongkol, Minh Triet Tran, Tin Trong Lu, John Darwin Ruanto, Zoe Black, Nanthinee Banditrattana, Boondarika Petersen, Jeremy Ross, Annette Sohn, Nittaya Phanuphak, Reshmie A. Ramautarsing

## Abstract

**Background:** Transgender people worldwide are disproportionately affected by physical and mental health conditions. Systemic marginalization, stigma, discrimination, and violence contribute to their adverse health outcomes. Little is known about these factors, or their impact on health and wellbeing, among transgender people in South East Asia.

**Methods:** The South East Asian Transgender Health Cohort (SEATrans), clinicaltrial.gov registration number NCT05454579) is the first multisite, prospective observational cohort study for transgender people in the region, including participants from Thailand, The Philippines and Vietnam. The study aims to track physical and mental health; identify biomedical, structural, and psychosocial factors impacting physical and mental health; describe structural barriers to healthcare; and develop guidance on holistic healthcare policies for transgender people in South East Asia. The study will enroll and follow-up 450 transgender people (300 HIV-negative people, 150 people living with HIV) from four community health centers. Sociodemographic information, behavioral risk information, mental health information (e.g., depression, anxiety, post-traumatic stress disorder), and data on psychosocial barriers will be collected. Laboratory testing will include testing for HIV, syphilis, *Chlamydia trachomatis*, *Neisseria gonorrhea*, hepatitis B and C, metabolic profile (glucose, cholesterol, HDL, LDL, triglycerides), and hormone levels. Participants will be followed until up to month 24. Participants with physical and/or mental health diagnoses will receive treatment or be linked to care. Descriptive analyses will be conducted to summarize sociodemographic characteristics, physical and mental health status, and structural and psychosocial factors, while regression analyses will assess relationships between several variables and outcomes of interest.

**Expected Impact:** Data from this cohort will inform holistic healthcare policies for transgender people in Thailand, The Philippines and Vietnam, as well as multilevel interventions to address the risks faced by this population. Tailored service packages will combine HIV, sexual, psychosocial health and gender-affirming care.

## Introduction

Transgender people are persons whose gender identity differs from the gender that was assigned to them at birth (1). Worldwide, transgender women are especially disproportionately affected by HIV. Their risk of HIV infection is almost 49 times higher than the risk to members of the general population (2). Furthermore, compared to cisgender people living with HIV (PLHIV), transgender women living with HIV are less likely to receive antiretroviral therapy (ART) and have higher rates of HIV-related mortality, whereas those who are receiving ART are more likely to have lower adherence and detectable viral load. Moreover, among HIV-negative transgender women, the uptake of and adherence to pre-exposure prophylaxis (PrEP) is lower compared to other populations (3–10). In addition, although many studies have raised concerns that transgender men also are at higher risk for HIV, particularly among those who have sex with cisgender men who have sex with men, the sexual health of transgender men remains understudied (11).

Thailand, The Philippines and Vietnam are also experiencing an HIV epidemic among transgender women. HIV prevalence among transgender women in these three countries ranges from 11.8% to 18% (12, 13), considerably higher than the prevalence in the general population (0.9%) (14). In Thailand, HIV prevalence and incidence have remained high among transgender women despite the introduction of highly effective HIV prevention measures such as treatment as prevention and PrEP (15). In the Philippines, the few studies that have assessed HIV epidemiology among transgender women indicate a prevalence of up to 11.4%, while other research suggests that 48% to 79% of transgender women had never been tested for HIV (16, 17). A recent report from Vietnam using respondent driven sampling found an HIV prevalence of 16.5% among transgender women (18). HIV prevalence data on transgender men are not available from these settings. Overall, very little evidence to guide the engagement of transgender people in HIV prevention and care are available from most Asian countries.

In addition to HIV, transgender people experience a high prevalence of several other adverse health outcomes, such as cardiovascular disease and mental health problems (14, 19). Important contributors to the HIV and other health vulnerability of transgender people include biomedical risks such sexual and injection risks, and metabolic complications as a result of hormone therapy. Structural and psychosocial factors, such as barriers to health care, stigma and discrimination, substance use, and exposure to abuse and violence, are equally important drivers of the health disparities among transgender people (20–22). At the same time, the majority of transgender people across the world experience stigma and discrimination on a daily basis, including from healthcare providers, which discourages them from accessing traditional health care settings (23). Unfortunately, little is known about these factors in Asia, and how they might impact health outcomes and wellbeing among transgender people. The South East Asian Transgender Health Cohort (SEATrans) was established to better understand these factors among transgender people in South East Asia. It is the first multisite, prospective observational cohort study for transgender people in Thailand, The Philippines and Vietnam. It aims to assess the biomedical, structural and psychosocial risks among transgender people living with or without HIV. SEATrans will also monitor health outcomes including HIV seroconversion, PrEP uptake and adherence, ART uptake and adherence, viral suppression, and long-term metabolic complications. Data generated from this cohort will inform holistic health care policies for transgender people, as well as multi-level interventions to tackle the multi-level risks faced by this vulnerable population.

## Materials and Methods

### Aim, design and setting

SEATrans is a longitudinal observational cohort study that will prospectively collect and analyze data on the physical and mental health of transgender people in Thailand, The Philippines and Vietnam, as well as factors that impact physical and mental health among this population, such as psychosocial barriers. The study has been registered on clinicaltrials.gov under number NCT05454579.

The aims of SEATrans are to:

1. Longitudinally track physical and mental health
2. Identify biomedical, structural, and psychosocial factors impacting physical and mental health
3. Describe structural barriers to healthcare
4. Develop guidance on holistic healthcare policies

SEATrans aims to recruit a cohort of 450 transgender people from four community health centers in Thailand, The Philippines and Vietnam. Participants will be enrolled and followed for up to 24 months. Study sites are:

1. Rainbow Sky Association of Thailand (RSAT), Bangkok, Thailand: This community-based organization is situated in Ramkhamhaeng, a highly populated suburban area of Bangkok with Thailand’s largest public university. It serves men who have sex with men and transgender women and provides a range of HIV and related health services as part of the Key Population-led Health Services Model, such as PrEP, testing for sexually transmitted infections, and same day initiation of antiretroviral therapy (24–27).
2. Tangerine Clinic, Bangkok, Thailand: Located in central Bangkok, Tangerine clinic is a transgender-led clinic, which has implemented an integrated gender-affirming and sexual health service model since its establishment in 2015. Tangerine clinic has provided services to almost 6,000 transgender women and men (28).
3. Glink Clinics, Ho Chi Minh City (HCMC), Vietnam: A community clinic in HCMC, offering a wide range of services to men who have sex with men and transgender women, including HIV related services, mental healthcare, and transgender healthcare, including hormone services, gender affirming surgery consultations and sexual health care.
4. Victoria by LoveYourself, Manila, the Philippines: A community clinic in Manila, The Philippines, and the first community clinic that offers services focused on the needs of transgender people in The Philippines. In addition to HIV screening and counseling services, hormone management and assessment and counseling in regards to gender affirming surgery are offered.

### Sample size

The sample size was calculated based on the prevalence of physical and mental health of transgender women. Between 2019 and 2022, the prevalence of syphilis, *Chlamydia trachomatis* (CT), and *Neisseria gonorrhea* (NG) in the Tangerine Clinic, Bangkok, Thailand were 6.3%, 20%, and 14.5%, respectively, while depression prevalence was 23%, 12% had anxiety symptoms, and 13% had post-traumatic stress disorder (PTSD) (29). Accounting for a lost to follow-up rate of 20%, the estimated sample size needed to assess these health outcomes is a total of 420 transgender people.

Furthermore, transgender women living with HIV have a higher prevalence of syphilis, CT, NG, depression and PTSD compared to transgender women living without HIV (29–31). Therefore, participants living with HIV will be enrolled in a ratio of 1:2 to those living without HIV. The number of participants calculated is rounded up to 450 (300 HIV-negative transgender people and 150 transgender people living with HIV), as follows:

1. 300 HIV negative transgender people (150 of whom are using or will start to use PrEP, and 150 who are not using and not willing to use PrEP)

- 190 Thai participants
- 70 Filipino participants
- 40 Vietnamese participants.
2. 150 transgender people living with HIV (either known or newly diagnosed)

- 90 Thai participants
- 40 Filipino participants
- 20 Vietnamese participants.

### Procedures

Also see tables 1-3 for study procedures by study visit.

**Table 1.** Study Visits and Procedures for Participants Living with HIV.

| Procedures | Screening | Baseline<sup>1</sup> | Month 6 | Month 12 | Month 18 | Month 24 |
| --- | --- | --- | --- | --- | --- | --- |
| Informed consent | X |  |  |  |  |  |
| Sociodemographic information |  | X |  |  |  |  |
| Vital signs (body temperature, blood pressure, pulse rate), body weight, height |  | X | X | X | X | X |
| Questionnaires<br>Behavioral risk assessment<br>Mental health assessment (screening for depression, anxiety, and PTSD)<br>Psychosocial questionnaire |  | X | X | X | X | X |
| Assessment of adherence to ART<sup>2</sup> |  | X | X | X | X | X |
| Risk reduction counseling and provision of condoms and lubricant<sup>2</sup> |  | X | X | X | X | X |
| Anti-HIV (3 ml) |  | X |  |  |  |  |
| Viral load (6 ml) |  | X | X | X | X | X |
| CD4 count (2 ml) |  | X |  | X |  | X |
| HBsAg (3 ml) |  | X |  |  |  |  |
| Anti-HBs (3 ml) |  | X |  |  |  |  |
| Blood Glucose (2 ml) |  | X | X | X | X | X |
| Lipid profile (Cholesterol, HDL, LDL, and Triglyceride) (3 ml) |  | X | X | X | X | X |
| Hormone levels (Estradiol and/or Testosterone) (1 ml) |  | X | X | X | X | X |
| Syphilis (3 ml) |  | X | X | X | X | X |
| CT/NG |  | X | X | X | X | X |
| Anti-HCV (3 ml) |  | X | X | X | X | X |
| HCV-RNA (HCV-VL and HCV-Genotype)<sup>3</sup> (6 ml) |  | X | X | X | X | X |
| Maximum blood volume required per visit (ml) | 0 | 35 | 24 | 26 | 24 | 26 |

Abbreviations: PTSD – post traumatic stress disorder, ART – antiretroviral therapy, HBsAg – hepatitis B surface antigen, anti-HBs – hepatitis B surface antibody, HDL – high density lipoprotein, LDL – low density lipoprotein, CT – Chlamydia Trachomatis, NG – Neisseria Gonorrhea, HCV – hepatitis C
<sup>1</sup> The baseline visit can happen on the same day as the screening.
<sup>2</sup> These are conducted at all sites as standard of care
<sup>3</sup> HCV-VL testing will be performed on participants who test reactive/positive for anti-HCV. HCV-Genotype testing will be performed for participants with an anti-HCV reactive/positive result and adequate HCV viral load, if applicable. If HCV-Genotype testing is not available, the samples will be stored until funding becomes available

**Table 2.** Study Visits and Procedures for HIV-Negative Participants who Already Received PrEP or Accept PrEP.

| Procedures | Screening | Baseline <sup>1</sup> | Month 1 | Month 3 | Month 6 | Month 9 | Month 12 | Month 15 | Month 18 | Month 21 | Month 24 |
| --- | --- | --- | --- | --- | --- | --- | --- | --- | --- | --- | --- |
| Informed consent | x |  |  |  |  |  |  |  |  |  |  |
| Sociodemographic information |  | x |  |  |  |  |  |  |  |  |  |
| Vital signs (body temperature, blood pressure, pulse rate), body weight, height |  | x | x | x | x | x | x | x | x | x | x |
| Questionnaires<br>Behavioral risk assessment<br>Mental health assessment (screening for depression, anxiety, and PTSD)<br>Psychosocial questionnaire |  | x |  |  | x |  | x |  | x |  | x |
| Assessment of adherence to PrEP<sup>2</sup> |  | x | x | x | x | x | x | x | x | x | x |
| Risk reduction counseling and provision of condoms and lubricant<sup>2</sup> |  | x | x | x | x | x | x | x | x | x | x |
| Anti-HIV (3 ml) |  | x | x | x | x | x | x | x | x | x | x |
| Creatinine (3 ml) |  | x |  |  | x |  | x |  | x |  | x |
| HBsAg (3 ml) |  | x |  |  |  |  |  |  |  |  |  |
| Anti-HBs (3 ml) |  | x |  |  |  |  |  |  |  |  |  |
| Blood Glucose (2 ml) |  | x |  |  | x |  | x |  | x |  | x |
| Lipid profile (Cholesterol, HDL, LDL, and Triglyceride) (3 ml) |  | x |  |  | x |  | x |  | x |  | x |
| Hormone levels (Estradiol and/or Testosterone) (1 ml) |  | x |  |  | x |  | x |  | x |  | x |
| Syphilis (3 ml) |  | x |  |  | x |  | x |  | x |  | x |
| CT/NG |  | x |  |  | x |  | x |  | x |  | x |
| Anti-HCV (3 ml) |  | x |  |  | x |  | x |  | x |  | x |
| HCV-RNA (HCV-VL and HCV-Genotype)<sup>3</sup> (6 ml) |  | x |  |  | x |  | x |  | x |  | x |
| Maximum blood volume required per visit (ml) | 0 | 30 | 3 | 3 | 24 | 3 | 24 | 3 | 24 | 3 | 24 |

Abbreviations: PrEP – pre-exposure prophylaxis, PTSD – post traumatic stress disorder, HBsAg – hepatitis B surface antigen, anti-HBs – hepatitis B surface antibody, HDL – high density lipoprotein, LDL – low density lipoprotein, CT – Chlamydia Trachomatis, NG – Neisseria Gonorrhea, HCV – hepatitis C
<sup>1</sup> The baseline visit can happen on the same day as the screening.
<sup>2</sup> These are conducted at all sites as the standard of care
<sup>3</sup> HCV-VL testing will be performed on participants who test reactive/positive for anti-HCV. HCV-Genotype testing will be performed for participants with an anti-HCV reactive/positive result and adequate HCV viral load, if applicable. If HCV-Genotype testing is not available, the samples will be stored until funding becomes available

**Table 3.** Study Visits and Procedures for HIV-negative Participants who Decline PrEP.

| Procedures | Screening | Baseline<sup>1</sup> | Month 3 | Month 6 | Month 9 | Month 12 | Month 15 | Month 18 | Month 21 | Month 24 |
| --- | --- | --- | --- | --- | --- | --- | --- | --- | --- | --- |
| Informed consent | X |  |  |  |  |  |  |  |  |  |
| Sociodemographic information |  | X |  |  |  |  |  |  |  |  |
| Vital signs (body temperature, blood pressure, pulse rate), body weight, height |  | X | X | X | X | X | X | X | X | X |
| Questionnaires<br>Behavioral risk assessment<br>Mental health assessment (screening for depression, anxiety, and PTSD)<br>Psychosocial questionnaire |  | X |  | X |  | X |  | X |  | X |
| Risk reduction counseling and provision of condoms and lubricant<sup>2</sup> |  | X | X | X | X | X | X | X | X | X |
| Anti-HIV (3 ml) |  | X | X | X | X | X | X | X | X | X |
| HBsAg (3 ml) |  | X |  |  |  |  |  |  |  |  |
| Anti-HBs (3 ml) |  | X |  |  |  |  |  |  |  |  |
| Blood Glucose (2 ml) |  | X |  | X |  | X |  | X |  | X |
| Lipid profile (Cholesterol, HDL, LDL, and Triglyceride) (3 ml) |  | X |  | X |  | X |  | X |  | X |
| Hormone levels (Estradiol and/or Testosterone) (1 ml) |  | X |  | X |  | X |  | X |  | X |
| Syphilis (3 ml) |  | X |  | X |  | X |  | X |  | X |
| CT/NG |  | X |  | X |  | X |  | X |  | X |
| Anti-HCV (3 ml) |  | X |  | X |  | X |  | X |  | X |
| HCV-RNA (HCV-VL and HCV-Genotype)<sup>3</sup> (6 ml) |  | X |  | X |  | X |  | X |  | X |
| Maximum blood volume required per visit (ml) | 0 | 27 | 3 | 21 | 3 | 21 | 3 | 21 | 3 | 21 |

Abbreviations: PrEP – pre-exposure prophylaxis, PTSD – post traumatic stress disorder, HBsAg – hepatitis B surface antigen, anti-HBs – hepatitis B surface antibody, HDL – high density lipoprotein, LDL – low density lipoprotein, CT – Chlamydia Trachomatis, NG – Neisseria Gonorrhea, HCV – hepatitis C
<sup>1</sup> The baseline visit can happen on the same day as the screening.
<sup>2</sup> These are conducted at all sites as the standard of care
<sup>3</sup> HCV-VL testing will be performed on participants who test reactive/positive for anti-HCV. HCV-Genotype testing will be performed for participants with an anti-HCV reactive/positive result and adequate HCV viral load, if applicable. If HCV-Genotype testing is not available, the samples will be stored until funding becomes available

#### Eligibility and recruitment

People are eligible to participate in the study if they:

- Are of Thai, Filipino, or Vietnamese nationality
- Are aged ≥ 18 years old
- Self-identify as a transgender woman or man
- Have provided informed consent.

Eligible transgender people will be asked by site staff about their interest in participating in the study. Once a participant is enrolled in the cohort study, they will receive a reminder of their upcoming visit by their preferred contact method. For the design of additional retention strategies, consultations with the community at each site will be held.

#### Informed consent

A trained member of the study staff will inform the participant of all aspects of the study using the national language or languages understandable to the participants at each site. Written information approved by the local Institutional Review Board (IRB) will be used. The study staff will ensure that before the participants decide to participate in the study, they understand its purpose, the procedures, and any risks or benefits associated with the study, and that taking part in the study is voluntary. A copy of the consent form will be given to the participant to keep. The consent process will be conducted in a private room. No study procedures will occur before the participant gives informed consent.

#### Sociodemographic information

Sociodemographic information will be collected by study staff at baseline. Information collected will include date of birth, nationality, sex at birth, gender identity, relationship status, education, housing situation, employment status, type of employment (including sex work), income, and number of people dependent on participant’s income.

#### Medical information

Medical information such as medical history and history of gender affirming procedures will be collected from the participants by study staff.

#### Vital signs and measurements

Body temperature, height, body weight, blood pressure, and pulse rate will be measured and recorded, and body mass index (BMI) will be calculated.

#### Questionnaires

Participants will be asked to complete three questionnaires:

1. Behavioral risk assessment (assessing risk behavior, including substance use).
2. Mental health assessment (screening for depression, anxiety, and PTSD). Depression screening will be conducted by patient health questionnaire 9 (PHQ-9) (32), anxiety screening by general anxiety disorder 7 (GAD-7) anxiety scale (33), and PTSD screening will be conducted by the PTSD checklist for DSM-5 (PCL-5) (34).
3. Psychosocial questionnaire (exploring experiences with stigma and discrimination; wellbeing; suicidal ideation and attempts; gender-based violence, etc).

The three sets of questionnaires will be all self-administered. If a participant cannot read due to a disability, and has given consent in the consent form to allow study staff to assist with the questionnaire, a designated study staff will read the questions out loud for the participant in a private room. The participant can then choose an answer, and the study staff will record the answers.

#### Physical Health

Laboratory tests will either be conducted at the study site, or samples will be collected and sent to affiliated laboratories for testing. The following laboratory tests will be conducted to assess a participant’s physical health:

1. HIV testing Participants will receive an HIV-test at the baseline visit to identify the current HIV status. In addition, HIV testing will be performed for HIV-negative participants at every visits and PrEP will be offered to all HIV-negative participants. Both laboratory-based methods and self-test method are acceptable for HIV testing. Either oral fluid-based or blood-based sample can be used for HIV self-test. If the self-testing result is reactive, a laboratory-based test is required to confirm the result.
2. Syphilis testing Blood samples will be collected for syphilis testing using the treponemal-based reverse algorithm. If the treponemal-based test is reactive, non-treponemal tests with titers will be performed. The non-treponemal-based traditional algorithm can also be performed prior to treponemal test confirmation. If the participant has previously been diagnosed with syphilis, treponemal testing is optional while non-treponemal testing is required.
3. NG and CT testing Any of pharyngeal swab, rectal swab, vaginal swab, neovaginal swab, urethral swab, or urine will be collected. Either separated or pooled samples can be used as a sample for testing.
4. Hepatitis B virus (HBV) and Hepatitis C virus (HCV) testing Blood samples will be collected for HBsAg, Anti-HBs, and Anti-HCV testing. HCV-VL testing will be performed on participants who test reactive for anti-HCV. At sites where HCV genotyping is available, this test will be performed on participants with a positive anti-HCV result and adequate HCV viral load (>2000 IU/mL).
5. Other testing Additional laboratory testing will be conducted to assess metabolic health (blood glucose and lipid profile: cholesterol, HDL, LDL and triglycerides), hormone levels (estradiol and/or testosterone), and creatinine for participants on PrEP. People living with HIV will also receive HIV viral load and CD4 testing

### Standard of care at all sites

The following are not part of study procedures, but will be conducted as standard of care at all sites:

#### Risk Reduction Counseling and PrEP or ART adherence

Counseling, including risk-reduction counseling and discussion on PrEP or ART adherence for participants who receive PrEP or participants living with HIV, will be provided as appropriate according to the standard of care provided at each study site. Condoms and lubricants will be provided to all participants as needed. HIV-negative participants not using PrEP will be informed about and offered to start PrEP. Participants may request an extra visit outside of these scheduled testing if needed and can also access non-occupational post-exposure prophylaxis (nPEP) services if available at the study site.

#### Gender-affirming care

All sites offer gender affirming healthcare, such as hormone level monitoring, advise on gender affirming hormone use, and counseling in regards to gender affirming surgery. Referral to affiliated healthcare providers will be arranged as needed.

#### Prevention of HIV

Participants who are currently on PrEP or new PrEP users can access PrEP services offered at each study site.

#### Treatment of HIV

Participants who have been newly diagnosed with HIV at the baseline or any subsequent visits will be referred to initiate ART according to the standard of care at each study site. Similarly, known participants living with HIV will be referred to initiate ART according to the standard of care if they are not in care already.

#### Treatment of sexually transmitted infections (STIs)

Participants who have syphilis or CT/NG infection will be referred to receive treatment according to the standard of care at each study site.

#### Treatment of HCV

Participant who has been diagnosed with Hepatitis C infection will be referred to receive treatment according to the standard of care at each study site.

#### Treatment of Mental Health Diagnoses

Participants who are diagnosed with any mental health problem will be referred to see a physician, specialist, psychologist, or psychiatrist for treatment according to the standard of care.

### Study visits

#### Baseline visit

Screening and baseline visits can happen on the same day. After eligible participants provide informed consent, sociodemographic information will be collected, as well as biomedical information on health, gender affirmation, HIV status, cardiovascular health, and other general medical information. Vital signs will be measured (body temperature, blood pressure, pulse rate), as well as bodyweight and height. Participants will be asked to complete the behavioral risk assessment, mental health assessment, and psychosocial questionnaire. Risk reduction counseling will be conducted, and condoms and lubricants will be provided. People living with HIV will be asked about their adherence to ART. HIV negative people on PrEP will be asked about how they use it, while HIV negative people not on PrEP will be offered PrEP. Anti-HIV testing will be conducted, unless participants have pre-existing evidence of anti-HIV reactivity or evidence of current ART use. Reactive HIV-tests will be confirmed by laboratory-based testing. Other laboratory test that will be conducted are: HIV viral load and CD4 counts for people living with HIV, HBsAg, anti-HBs, glucose, lipid profile, hormone levels, syphilis testing, testing for CT and NG, anti-HCV (and HCV-RNA and HCV genotyping if anti-HCV is positive). Among participants on PrEP, creatinine will also be tested.

#### Follow-up visits for participants living with HIV

People living with HIV will be followed up every six months until study completion at month 24. The following will be conducted at every follow-up visit: measurement of vital signs and body weight, behavioral risk assessment, mental health assessment, psychosocial questionnaire, assessment of adherence to ART, risk reduction counseling, provision of condoms and lubricants, and laboratory assessment of HIV viral load, glucose, lipid profile, hormone levels, syphilis, CT/NG, anti-HCV (and HCV-RNA and HCV genotyping if anti-HCV is positive). CD4 count will be measured yearly (at months 12 and 24).

#### Follow-up visits for HIV-negative participants

HIV-negative participants will be followed-up every three months until study completion at month 24. The following will be conducted at each follow-up visit: measurement of vital signs and body weight, risk reduction counseling, provision of condoms and lubricants, assessment of PrEP use or offering of PrEP, and anti-HIV testing. All other procedures (behavioral risk assessment, mental health assessment, psychosocial questionnaire, creatinine testing for those using PrEP, glucose, lipid profile, hormone levels, syphilis, CT/NG, anti-HCV, and HCV-RNA and HCV genotyping if anti-HCV is positive), will be conducted every six months.

All variables including specimen collection to be collected will depend on participants’ willingness, clinical/physician’s decision, or per the standard service of the study sites. If participants visit the clinic between the schedules study visits, any data routinely gathered as part of the service will be collected. The visit window will be flexible to accommodate participants.

The use of laboratory results obtained from external laboratories within a validity period prior to each scheduled visit is allowed. If laboratory results from an external laboratory is available, laboratory testing at the site on the day of the study visit is optional. The following conditions apply for results from external laboratories:

1. For all laboratory testing results, the validity period is defined as three months or as determined by the physician, except HIV-negative and HIV-reactive results, reactive treponemal testing for syphilis, and anti-HCV positive results.
2. HIV-negative participants are considered to have a seven-day validity period for anti-HIV test results.
3. For people living with HIV, an HIV-reactive result obtained at any time before enrollment or each follow up visit or any evidence of current ART can be used to confirm HIV status. Anti-HIV testing at the scheduled visit is not required to confirm the HIV status.
4. Reactive treponemal results for syphilis testing obtained at any time before enrollment or each follow up visit can be used. If the previous results are not available, the clinical history of syphilis must be documented.
5. The anti-HCV positive results that obtained at any time before enrollment or each follow up visit can be used. Anti-HCV testing is not required to confirm the HCV status.

### Data collection and management

An overview of the data collected for the study is outlined in Table 4.

**Table 4.** Outcome data collected in SEATrans.

| Category | Method | Details | Frequency |
| --- | --- | --- | --- |
| Sociodemographic information | Questions asked by study staff | Gender identity | Once, at baseline |
|  |  | Sex assigned at birth |  |
|  |  | Age |  |
|  |  | Education level |  |
|  |  | Sex work |  |
|  |  | Housing |  |
| Measurements | Physical examination | Height | Once, at baseline |
|  |  | Pulse rate | At every visit |
|  |  | Blood pressure |  |
|  |  | Body temperature |  |
|  |  | Body weight |  |
| Medical history | Questions asked by study staff | Underlying conditions | At every visit |
|  |  | Medication use |  |
| Gender affirmation | Questions asked by study staff | Age of awareness of trans identity | Once, at baseline |
|  |  | Age of gender expression |  |
|  |  | Hormone use | At every visit |
|  |  | Surgery status |  |
|  |  | Silicone injections |  |
| Behavioral risk assessment | Self-administered questionnaire | Number and type of sexual partners | At baseline and every 6 months |
|  |  | Condom use |  |
|  |  | Substance use |  |
| Mental health assessment | Self-administered questionnaire | Depression | At baseline and every 6 months |
|  |  | Anxiety |  |
|  |  | PTSD |  |
| Psychosocial questionnaire | Self-administered questionnaire | Experience of stigma and discrimination | At baseline and every 6 months |
|  |  | Well-being |  |
|  |  | Suicidal ideation and attempts |  |
|  |  | Gender-based violence |  |
| Clinical information – HIV related | Laboratory testing | HIV test results | At baseline and every 3 months |
|  |  | Viral load | At baseline and every 6 months |
|  |  | CD4 count | At baseline and every 12 months |
|  | Questions asked by study staff | ART use | At every visit |
|  |  | PrEP use | At every visit |
| Clinical information – STI related | Laboratory testing | Syphilis | At baseline and every 6 months |
|  |  | NG |  |
|  |  | CT |  |
|  |  | HCV |  |
| Clinical information – others | Laboratory testing | HBV | At baseline |
|  |  | Blood glucose | At baseline and every 6 months |
|  |  | Lipid profile (cholesterol, HDL, LDL and triglycerides) |  |
|  |  | Hormone levels (estradiol and/or testosterone) |  |
|  |  | Creatinine |  |

Abbreviations: PTSD – post-traumatic stress disorder, ART – antiretroviral therapy, PrEP – pre-exposure prophylaxis, NG – Neisseria Gonorrhea, CT – Chlamydia Trachomatis, HCV – hepatitis C virus, HBV – hepatitis B virus

Study data will be collected and managed using REDCap electronic data capture tools hosted at The Institute of HIV Research and Innovation (IHRI) (35, 36). REDCap (Research Electronic Data Capture) is a secure, web-based software platform designed to support data capture for research studies. In addition, IHRI will provide regulatory support and monitoring, site compliance with study procedures, and any technical assistance to participating sites as needed. All sites will be able to enter data in their site-specific eCRFs, and individual sites will not be able to access eCRFs from other sites.

Once sites complete and submit their eCRF, all data will be stored on a central server at IHRI. From this point, only the IHRI data management team and the monitoring team will have access to the data. IHRI will maintain the REDCap database, review data for completeness and legibility, and coordinate with study sites to resolve any outstanding queries. For analysis and preparation of reports and publications, data can be shared securely with other members of the study teams (e.g., investigators, statisticians).

### Safety considerations

SEATrans is an observational cohort study, and does not meet the criteria of a clinical trial. Therefore, a Data Safety Monitoring Board is not required. Nonetheless, adverse events can occur (such as injuries, psychological or social harms), and will be reported to the IRBs.

Participants may feel uncomfortable or discouraged when asked to complete questionnaires about risky behavior or disclose confidential information. The participants; identification information records will be kept separate from the research data and stored in a lockable filing cabinet to ensure anonymity of the data. In addition, the electronic data capture will store participant information on a secure computer network.

The risk of primary concern in this study is related to ascertaining and providing HIV diagnostic information and, in particular, involuntary disclosure of HIV status to others. These disclosures may result in depression and rarely suicide among individuals learning that they have HIV. Furthermore, involuntary disclosure to others may result in extreme prejudice by the community, family, employers, and psychosocial factors including stigma and discrimination. This risk will be minimized by the fact that all counselors will be trained in pre-and post-test counseling for HIV and will aim at fully informing participants of all activities in the study and attendant risks and benefits. Risks associated with HIV diagnosis will be minimized by providing information regarding prognosis, transmissibility, and potential steps which can be taken to extend life through opportunistic infection management, prevention of transmission of HIV, and seeking additional support and care. Further, should the participants’ HIV test results become known to others, the study staff will take appropriate action to assist participants with any discrimination they may experience during their participation in this study. Such measures will be accompanied by appropriate counseling. However, the study staff will take great care to minimize this risk occurring. To ensure this, personally identifying information will be collected at the time of enrollment and stored in a secure, locked cabinet, to which only key study team members will have access. Participants will be allocated a participant identification number, which will be used on all subsequent visits. This measure will ensure confidentiality of personal information and minimize the chances of their HIV status, and other participant information becoming known to others.

There may be risks associated with the collection of anal and urethral/neovaginal specimens for STI screening and diagnosis. Participants may feel uncomfortable and shy about having the specimens collected. The swab used to collect specimens may cause some irritation, but it will not cause pain. The anal and urethral/neovaginal collection will be done by a well-trained staff or self-collection in a private room. Every effort will be made to make sure that the participant feels comfortable during the procedure.

### Type of data and statistical analyses planned

Descriptive analyses will summarize sociodemographic characteristics, physical and mental health status, and structural and psychosocial factors. These data will be presented as mean and standard deviation or median and interquartile range (IQR) for continuous variables. Categorical variables will be presented as proportions. For comparisons among groups, e.g. health status or structural and psychosocial factors among transgender people living with HIV compared to those living without HIV, the student’s t-test will be used for the mean comparison of a continuous variable with normal distribution among two groups. The median test will be used for the non-normally distribution continuous variables. To determine relationships between categorical variables, chi-squared tests will be used. Prevalence and incidence of physical and mental health issues will be presented with a 95% confidence interval (CI). Any factors impacting physical health (e.g. HIV, syphilis, CT, and NG) will be analyzed using logistic regression analysis. In addition, linear regression and/or logistic regression will be used to determine factors associated with mental health problems.

### Ethical considerations and declarations

This study will be conducted in compliance with the International Conference on Harmonization (ICH) guidelines for Good Clinical Practice (GCP) and with the guidelines of the World Medical Assembly Declaration of Helsinki. The protocol, informed consent form, data collection forms, and the questionnaires have been reviewed and approved by the relevant IRBs or Ethics Committees as follows: IRB of the Faculty of Medicine, Chulalongkorn University, Bangkok, Thailand on 25 August 2022; The Research Ethics Committee of University of Medicine and Pharmacy at Ho Chi Minh City, Vietnam on 14 December 2023; and the National Ethics Committee, Philippine National Health Research System, The Philippines on 5 April 2024. Participants will provide written informed consent before any study-related procedures are conducted, and will be re-consented if needed due to protocol changes.

### Status and timeline of the study

SEATrans started recruitment on 2 December 2022. As of March 2025, 450 transgender people with a median (IQR) age of 28.0 (23.0-33.0) years were enrolled and completed their baseline visit. Of these, 414 (92.0%) were transgender women, 33 (7.3%) were transgender men, and 3 (0.7%) identified as non-binary; 280 (62.2%) were Thai, 110 (24.4%) were Filipino, and 60 (13.3%) were Vietnamese. One hundred and fifty (33.3%) were living with HIV, while 150 (50.0%%) of 300 HIV-negative participants were on PrEP. The study closed on 30 April 2025.

## Discussion

SEATrans was planned as the first observational cohort study of transgender persons in South East Asia to assess physical and mental health, and to explore the impact of social and structural barriers on health. All 450 participants have been enrolled, 315 participants completed their month 6 visit, 199 completed month 12, 108 completed month 18, and 39 completed the full follow-up of 24 months.

Potential limitations of the study are that findings from SEATrans are potentially not generalizable to transgender populations in settings outside of Thailand, The Philippines and Vietnam, or to transgender populations in more rural settings in these countries. Furthermore, only a small proportion of participants identified as non-binary, and more research is needed to explore health and wellbeing among this population. Finally, participating in a longitudinal cohort study can be demanding for participants and staff of the study sites. To minimize this burden, visit windows were flexible, and laboratory results from external laboratories could be used for the study.

All findings will be disseminated broadly through several methods. Findings will first and foremost be shared with affected and participating communities. Findings will also be shared at academic conferences; at national, regional and international meetings, and in peer-reviewed journals.

In summary, SEATrans is the first observational cohort study to explore physical and mental health, and social determinants of health among transgender people in South East Asia. Findings will elucidate the burden of health issues and psychosocial issues among this population, the interrelatedness of these issues, and guide recommendations to strengthen competent services and enable systems for health for transgender people in South East Asia.

## Authors’ contributions

RJ, RAR, and NP conceptualized the study. RJ and RAR drafted the outline of the manuscript. RJ, TP, SS, TL, RP, PP, TA, VS, NW, TS, KM, MTT, TTL, JDR, ZB, NB, BP, JR, AS, NP and RAR reviewed and edited the draft.

## Acknowledgements

We are grateful to all participants in the SEATrans cohort, all providers delivering care at the participating community clinics, and all study staff.

## Data availability

All relevant data from this study will be made available upon reasonable request to the authors

## References

1. Standards of care for the health of transsexual, transgender and gender nonconforming people. Minneapolis: World Professional Association for Transgender Health 2012.

2. Baral SD, Poteat T, Stromdahl S, Wirtz AL, Guadamuz TE, Beyrer C. Worldwide burden of HIV in transgender women: a systematic review and meta-analysis. Lancet Infect Dis. 2013;13(3):214–22.

3. Deutsch MB, Glidden DV, Sevelius J, Keatley J, McMahan V, Guanira J, et al. HIV pre-exposure prophylaxis in transgender women: a subgroup analysis of the iPrEx trial. Lancet HIV. 2015;2(12):e512–9.

4. Health SFDoP. HIV/AIDS Epidemiology Annual Report. 2008.

5. Melendez RM, Exner TA, Ehrhardt AA, Dodge B, Remien RH, Rotheram-Borus MJ, et al. Health and health care among male-to-female transgender persons who are HIV positive. Am J Public Health. 2006;96(6):1034–7.

6. Sevelius JM, Carrico A, Johnson MO. Antiretroviral therapy adherence among transgender women living with HIV. J Assoc Nurses AIDS Care. 2010;21(3):256–64.

7. Reisner SL, White Hughto JM, Pardee D, Sevelius J. Syndemics and gender affirmation: HIV sexual risk in female-to-male trans masculine adults reporting sexual contact with cisgender males. Int J STD AIDS. 2016;27(11):955–66.

8. Sevelius JM, Keatley J, Calma N, Arnold E. ’I am not a man’: Trans-specific barriers and facilitators to PrEP acceptability among transgender women. Glob Public Health. 2016;11(7-8):1060–75.

9. Wilson EC, Jin H, Liu A, Raymond HF. Knowledge, Indications and Willingness to Take Pre-Exposure Prophylaxis among Transwomen in San Francisco, 2013. PLoS One. 2015;10(6):e0128971.

10. Ramautarsing RA, Meksena R, Sungsing T, Chinbunchorn T, Sangprasert T, Fungfoosri O, et al. Evaluation of a pre-exposure prophylaxis programme for men who have sex with men and transgender women in Thailand: learning through the HIV prevention cascade lens. J Int AIDS Soc. 2020;23 Suppl 3:e25540.

11. Kenagy GP, Hsieh CM. The risk less known: female-to-male transgender persons’ vulnerability to HIV infection. AIDS care. 2005;17(2):195–207.

12. HIV prevalence among transgender people (male-to-female), 2019 [Internet]. 2019. Available from: https://www.aidsdatahub.org/Key-Populations/Transgender-people.

13. Colby D, Nguyen NA, Le B, Toan T, Thien DD, Huyen HT, et al. HIV and Syphilis Prevalence Among Transgender Women in Ho Chi Minh City, Vietnam. AIDS and behavior. 2016;20(Suppl 3):379–85.

14. Reisner SL, Poteat T, Keatley J, Cabral M, Mothopeng T, Dunham E, et al. Global health burden and needs of transgender populations: a review. Lancet (London, England). 2016;388(10042):412–36.

15. van Griensven F, Phanuphak N, Manopaiboon C, Dunne EF, Colby DJ, Chaiphosri P, et al. HIV prevalence and incidence among men who have sex with men and transgender women in Bangkok, 2014-2018: Outcomes of a consensus development initiative. PLoS One. 2022;17(1):e0262694.

16. National Epidemiology Center, Department of Health, Philippines. IHBSS: Integrative HIV Behavioral and Serological Surveillance.; 2014.

17. World Health Organization. Policy brief: Transgender health and HIV in the Philippines.. 2016.

18. Vi VTT, Long KQ, Hong L, Anh HTN, Ngoc NV, Tam VV, et al. HIV Prevalence and Factors Related to HIV Infection Among Transgender Women in Vietnam: A Respondent Driven Sampling Approach. AIDS Behav. 2020;24(11):3132–41.

19. Cetlin M, Fulda ES, Chu SM, Hamnvik OR, Poteat T, Zanni MV, et al. Cardiovascular Disease Risk Among Transgender People with HIV. Curr HIV/AIDS Rep. 2021;18(5):407–23.

20. Baral S, Logie CH, Grosso A, Wirtz AL, Beyrer C. Modified social ecological model: a tool to guide the assessment of the risks and risk contexts of HIV epidemics. BMC Public Health. 2013;13:482.

21. Brennan J, Kuhns LM, Johnson AK, Belzer M, Wilson EC, Garofalo R, et al. Syndemic theory and HIV-related risk among young transgender women: the role of multiple, co-occurring health problems and social marginalization. Am J Public Health. 2012;102(9):1751–7.

22. Poteat T, Scheim A, Xavier J, Reisner S, Baral S. Global Epidemiology of HIV Infection and Related Syndemics Affecting Transgender People. J Acquir Immune Defic Syndr. 2016;72 Suppl 3:S210–9.

23. Winter S, Diamond M, Green J, Karasic D, Reed T, Whittle S, et al. Transgender people: health at the margins of society. Lancet (London, England). 2016;388(10042):390–400.

24. Differentiated HIV Service Delivery along the cascade for men who have sex with men and transgender women in Thailand: lessons learned from the LINKAGES project. 2018.

25. Lujintanon S, Amatavete S, Leenasirimakul P, Meechure J, Noopetch P, Sangtong S, et al. Acceptability and retention of the key population-led HIV treatment service for men who have sex with men and transgender women living with HIV in Thailand. J Int AIDS Soc. 2023;26(2):e26062.

26. Phanuphak N, Sungsing T, Jantarapakde J, Pengnonyang S, Trachunthong D, Mingkwanrungruang P, et al. Princess PrEP program: the first key population-led model to deliver pre-exposure prophylaxis to key populations by key populations in Thailand. Sex Health. 2018;15(6):542–55.

27. Thammajaruk N, Ramautarsing RA, Hiransuthikul A, Suriwong S, Tasomboon W, Thapwong P, et al. Pooled Pharyngeal, Rectal, and Urine Specimens for the Point-of-Care Detection of Chlamydia trachomatis and Neisseria gonorrhoeae by Lay Providers in Key Population-Led Health Services in Thailand. Pathogens. 2023;12(10).

28. Janamnuaysook R, Taesombat R, Wong J, Vannakit R, Mills S, van der Loeff MS, et al. Innovating healthcare: Tangerine Clinic’s role in implementing inclusive and equitable HIV care for transgender people in Thailand. J Int AIDS Soc. 2025;28(1):e26405.

29. Burden of PTSD, depression, and anxiety in transgender HIV+ and HIV – persons at key-population led health clinics in Bangkok and Pattaya: mixed methods cross-sectional screening study. Presented at the 10th IAS conference on HIV science (IAS 2019) – Mexico city, 21-24 July 2019. 2019.

30. Hiransuthikul A, Janamnuaysook R, Sungsing T, Jantarapakde J, Trachunthong D, Mills S, et al. High burden of chlamydia and gonorrhoea in pharyngeal, rectal and urethral sites among Thai transgender women: implications for anatomical site selection for the screening of STI. Sex Transm Infect. 2019;95(7):534–9.

31. van Griensven F, Janamnuaysook R, Nampaisan O, Peelay J, Samitpol K, Mills S, et al. Uptake of Primary Care Services and HIV and Syphilis Infection among Transgender Women attending the Tangerine Community Health Clinic, Bangkok, Thailand, 2016 - 2019. J Int AIDS Soc. 2021;24(6):e25683.

32. K K, RL S. The PHQ-9: A New Depression Diagnostic and Severity Measure. Psychiatric Annals. 2002;32(9):509–15.

33. Spitzer RL, Kroenke K, Williams JB, Lowe B. A brief measure for assessing generalized anxiety disorder: the GAD-7. Arch Intern Med. 2006;166(10):1092–7.

34. Blevins CA, Weathers FW, Davis MT, Witte TK, Domino JL. The Posttraumatic Stress Disorder Checklist for DSM-5 (PCL-5): Development and Initial Psychometric Evaluation. J Trauma Stress. 2015;28(6):489–98.

35. Harris PA, Taylor R, Minor BL, Elliott V, Fernandez M, O’Neal L, et al. The REDCap consortium: Building an international community of software platform partners. J Biomed Inform. 2019;95:103208.

36. Harris PA, Taylor R, Thielke R, Payne J, Gonzalez N, Conde JG. Research electronic data capture (REDCap)--a metadata-driven methodology and workflow process for providing translational research informatics support. J Biomed Inform. 2009;42(2):377–81.

